# Feasibility of Patient-Uploaded Videos for Gait Assessment in Multiple Sclerosis

**DOI:** 10.64898/2026.07.08.26356963

**Authors:** M McCune, Y Ackerman, A Camacho, N Sisodia, J Wijangco, K Henderson, J Bradsby, S Poole, A Torres-Espin, MJ Miller, VJ Block, R Bove

## Abstract

**Background:** Gait impairment is common among people with multiple sclerosis (PwMS) and is an important marker of disease progression. However, gait assessments typically require in-person evaluations.

**Objective:** To describe the pose-estimation-based method for estimating spatiotemporal gait parameters from a single consumer-grade video, and evaluate the feasibility of home video collection by PwMS.

**Methods:** In a single-center longitudinal digital phenotyping study, ambulatory adults with MS completed a standardized walking task recorded in the frontal plane. Pose estimation (MediaPipe Pose, Ultralytics) and custom scripts were used to estimate gait parameters from videos. Participants were invited to record walking videos at home using personal devices. Adoption and technical feasibility were evaluated across two home video data acquisition phases, with iterative protocol refinements.

**Results:** The in-clinic study included 132 participants; 55 contributed home videos. In Phase I, while home video adoption was low (45% [30/66]), 87% [26/30] uploaded ≥1 video of sufficient quality for gait analysis. After protocol refinements, 100% [25/25] uploaded ≥1 high-quality video. Overall, high-quality frontal-plane videos were obtained at similar rates at home (92% [97/105]) and in-clinic (91% [423/467]).

**Conclusions:** Home walking videos can feasibly be collected by PwMS to estimate gait parameters, providing an accessible approach for remote gait monitoring.

## INTRODUCTION

Multiple sclerosis (MS) is a chronic neurodegenerative condition, and the majority of people with MS (PwMS) have gait or balance impairments.^1,2^ Decreased walking speed, altered biomechanics, and fear of falling can all contribute to limited walking in community settings,^3^ negative impacts on social engagement and emotional health,^1,4^ and difficulty achieving the level of exercise recommended for PwMS.^5^

Given the significant impact of walking and mobility on quality of life, it is important to monitor gait changes early and often. 3D motion capture, force plates, and pressure mats are the gold standard tools for evaluating walking biomechanics and kinematics.^6^ However, these tools are not widely accessible given their high cost, the requirement for dedicated space, and the expertise required to interpret kinematic data. Furthermore, even for outcome measures with good psychometric properties and low burden, such as the Timed 25-Foot Walk (T25FW),^7^ in-person evaluations impose travel, scheduling, and time burdens on participants, limiting the frequency of these assessments.

Pose estimation software offers a potential solution for gait analysis from videos recorded with consumer-grade cameras, whether in-clinic or remotely. Pose estimation uses computer vision to automatically identify the position of body landmarks from color videos.^8,9^ Spatiotemporal gait parameters extracted from a single-color video using pose estimation in healthy controls and neurological patient populations have been validated against gold standard tools.^10,11^ However, previous work evaluating pose-estimation-derived gait parameters has typically been conducted in a controlled research setting with a defined walking task and cameras operated by the study team. We previously validated self-recorded videos as a feasible and valid method to collect dexterity measures using pose estimation.^12^ It remains unknown if spatiotemporal gait parameters can be estimated from walking videos recorded independently by PwMS at home.

To address this gap, this study aimed to 1) develop a pose-estimation-based method to estimate spatiotemporal gait parameters from in-clinic and home walking videos and 2) evaluate the feasibility of PwMS uploading home walking videos of sufficient quality for analysis using the pose estimation method. The overall strategy was to identify the video acquisition and processing strategy that imposes the least user burden.

## METHODS

### Study Setting and Participants

Participants were drawn from the ongoing BrainWalk digital phenotyping study at the University of California, San Francisco (UCSF). For the current analyses, adults (age ≥ 18 years) with a diagnosis of Clinically Isolated Syndrome (CIS)/MS by 2017 McDonald Criteria,^13^ and Expanded Disability Status Scale (EDSS)^14^ ≤ 6.5 were eligible. Exclusion criteria included other neurological or non-affective psychiatric disorders or insufficient visual, auditory, or motor capacity to complete tablet-based assessments (part of broader study assessments).

The UCSF Committee for Human Research approved all study procedures (IRB No. 21-33227). Written informed consent was obtained from all participants.

### Study Measures and Data Collection

Participants completed a baseline visit and annual follow-up visits. Gait was assessed using in-clinic and home video walking tasks.

#### In-Clinic Walking Tasks

Participants completed walking tasks on an instrumented walkway. The Zeno Walkway (ProtoKinetics LLC, Havertown, Pennsylvania) is a 16-foot instrumented walkway containing pressure sensors that utilize the ProtoKinetics Movement Analysis Software to generate ground-truth spatiotemporal gait parameters, with excellent reliability^15^ and concurrent validity against other industry standards.^16^ Participants completed multiple back-and-forth passes, turning off the walkway after each pass. Simultaneously, a single consumer-grade camera (Logitech C992) placed at one end of the walkway recorded frontal-plane videos of participants walking toward and away from the camera (**Figure 1A**).

**Figure 1.**
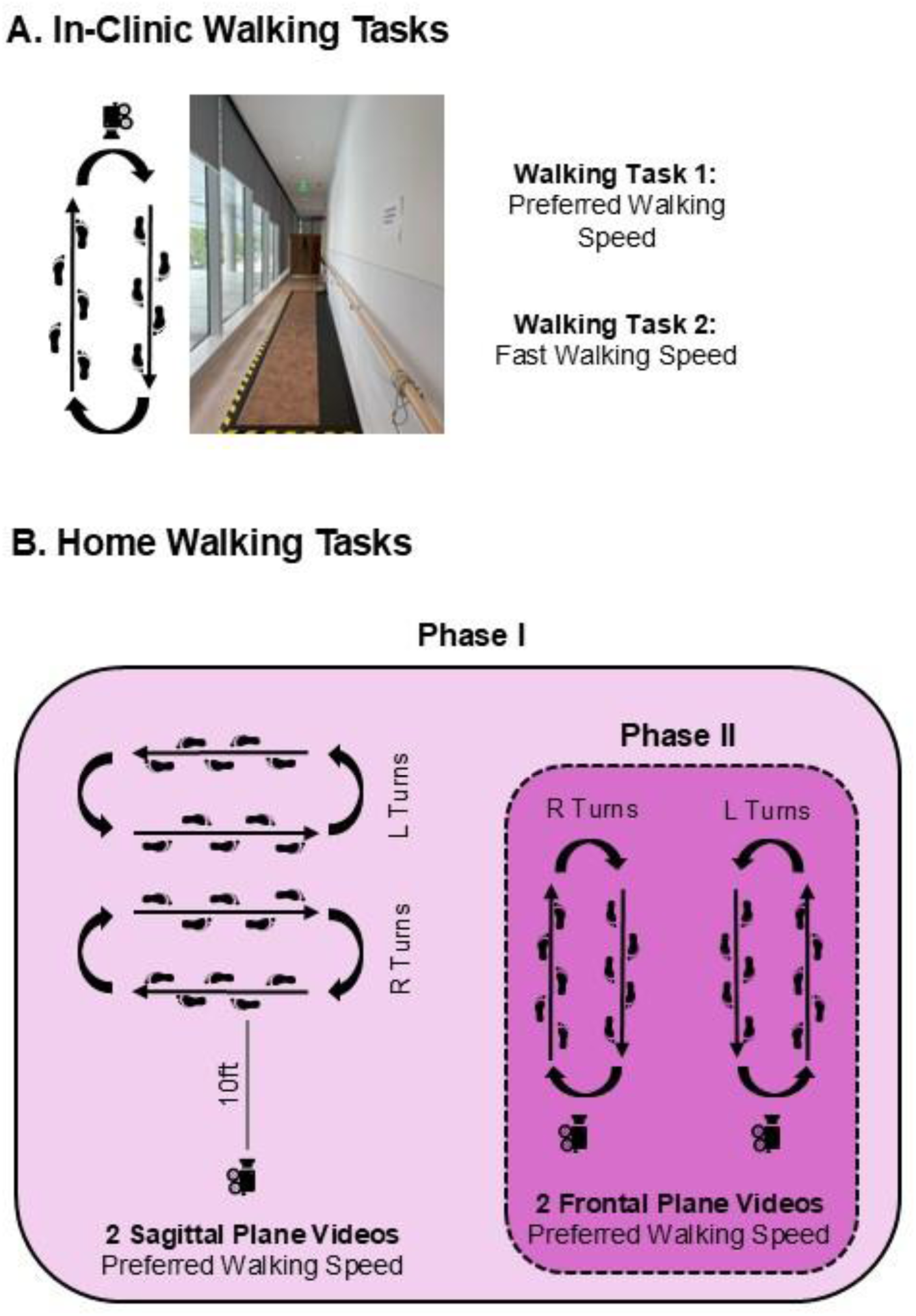
In-clinic and home walking tasks. A. In Clinic: During the in-clinic walking tasks, participants completed multiple back-and-forth passes on an instrumented walkway while simultaneously being recorded in the frontal plane by a single consumer-grade camera. B. Home: The home video walking tasks were similar to the in-clinic, with participants making multiple back-and-forth passes in front of the camera. In Phase I of home video data acquisition, participants recorded four walking videos at each time point: sagittal plane with right turns, sagittal plane with left turns, frontal plane with right turns, and frontal plane with left turns. In Phase II, participants were asked to record only the two frontal-plane videos.

Two walking tasks were analyzed: preferred walking speed (PWS), and fast walking (FW), each consisting of four walking passes with three turns.

#### Iterative Phases of Home Video Data Acquisition

Participants were invited to record videos of themselves walking in their homes with instructions provided electronically via email. Given the lack of prior data PwMS self-recording gait at home, the current study prioritized safety; therefore, participants were instructed to walk at a PWS that felt safe to them. No other specific criteria relating to ambulatory status were required.

The data acquisition protocol for home walking was iteratively refined, making improvements after reviewing common issues in Phase I videos. To summarize the major difference between the two Phases, in Phase I, participants sent videos in both the frontal and sagittal planes; in Phase II, participants sent only frontal-plane videos (**Figure 1B**).

### Video Processing Methods

This study aimed to minimize barriers to use by PwMS. To achieve this goal, the video processing method was designed to 1) only require a single video, avoiding the need for participants to own recording multiple devices, 2) automatically identify segments of the video that should analyzed and remove extraneous activity (i.e., camera setup), and 3) allow for videos with variations in recording environment (i.e., distance traveled, distance from camera to participant). Therefore, the gait parameters calculated in this project are not intended to be one-to-one substitutes for those collected using gold-standard tools such as 3D motion capture. Instead, the video processing methods summarized in **Figure 2** aimed to develop a set of gait parameters to provide a snapshot of patient gait, collected with consumer-grade tools across various settings.

**Figure 2.**
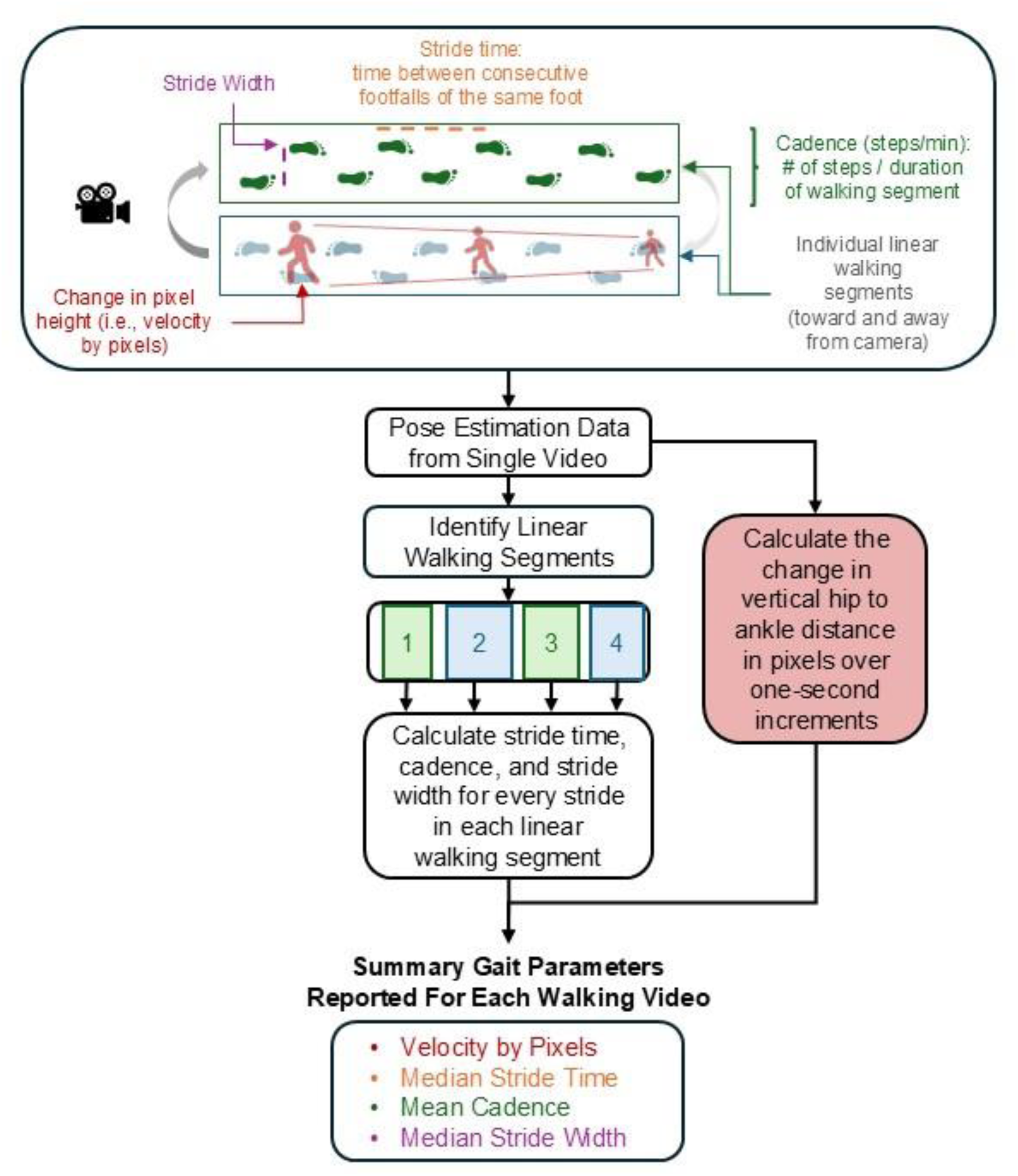
Schema describing gait parameters extracted from a single video. Velocity was estimated using the median change in pixel height per second (i.e., velocity by pixels), calculated across all one-second increments in the video that did not contain local minimums or maximums (potential turns) or gaps with missing landmark position data. Then the linear walking selection algorithm automatically identified segments of the pose estimation data that contained linear walking and sufficient marker visibility. Stride time, cadence, and stride width were calculated from each segment. The summary parameters in the lower box were calculated for each video.

#### Pose Estimation

A custom script was generated to integrate MediaPipe Pose and Ultralytics YOLOv8-pose pose estimation models. MediaPipe Pose and Ultralytics are open-access software packages that estimate the positions of body landmarks from RGB video frames.^8,17^ This custom script utilized the YOLOv8 model’s confidence estimate of whether the figure found in the image is a person and the MediaPipe Pose model’s estimate of how likely each landmark is visible in the frame. These two confidence estimates were particularly important for home videos, where it is common to have additional objects in the background (e.g., shadows, plants, furniture), and for some landmarks to be out of frame (e.g., feet and ankles when close to the camera).

The first processing step used the YOLOv8 confidence estimate that the figure is a person. If this estimate was ≥ 85%, a bounding box was created around the section of the image containing the person, and the 2D position of 17 body landmarks in pixels was output. Next, the MediaPipe Pose model was run on the section of the frame outlined by the YOLOv8 bounding box. The MediaPipe model then outputs the position of 33 landmarks in both 2D ‘pose’ units, defined as the landmark coordinates normalized to [0.0, 1.0] by the bounding box’s width and height, and ‘world’ coordinates, defined as real-world coordinates in meters with the origin at the midpoint between the two hip landmarks. Additionally, the model calculates a visibility score for each landmark at each frame, ranging from 0 to 1, with higher scores indicating a greater likelihood that the landmark is visible in the image.

This analysis utilized the left and right hip, ankle, and heel MediaPipe landmarks, along with their corresponding visibility scores, and the left and right hip and ankle YOLOv8 landmarks.

#### Automated Identification of Linear Walking

An algorithm was developed to automatically select segments of the raw pose estimation data in which the participant was walking toward or away from the camera with required lower-body landmarks visible (**Figure 3**, with further details in **Figure S1**). All videos were run through this rule-based algorithm to identify linear walking segments and exclude extraneous activities (e.g., camera setup, pauses) or frames with incomplete body capture.

**Figure 3.**
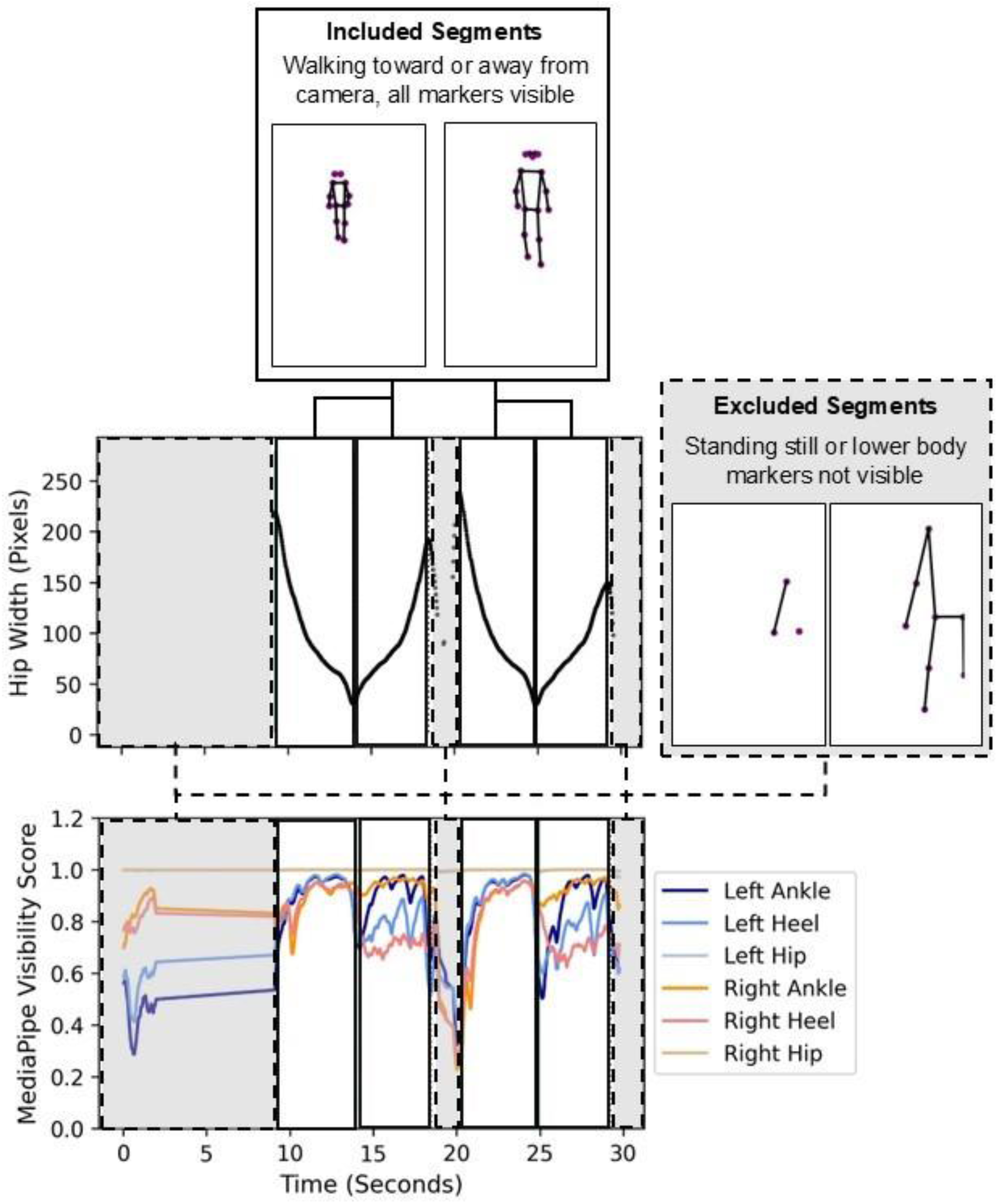
Overview of video segments that were included or excluded from an example video. Sections outlined in solid black were times in which the participant was walking toward or away from the camera with sufficient visibility of each marker. The sections outlined in dashed lines and highlighted in grey were excluded from the analysis due to poor marker visibility or walking pauses. Example UltralyticsYOLOv8-pose landmark positions at two included and two excluded frames are shown.

#### Spatiotemporal Gait Parameters from Pose Estimation Data

Within the linear walking segments, four gait parameters were calculated: a proxy gait velocity (pixel height per second, i.e., velocity by pixels), stride time, cadence, and stride width. These parameters are sensitive to MS-related gait impairment, as affected individuals walk more slowly, with longer stride times, decreased cadence, and a wider base of support.^18,19^ Except for the velocity by pixels, parameters were calculated for every stride (full calculation steps in **Table 1**, **Figures S2-S4**); mean and median values were summarized for each video (**Figure 2**).

**Table 1.** Spatiotemporal gait parameter definitions and inputs.

| Parameter Name | Parameter Definition | Inputs | Summary Statistics Per Video |
| --- | --- | --- | --- |
| **Velocity by Pixels** | Estimate of walking velocity, measured by the change in the distance between the hip and ankle landmarks over each one-second interval | Landmarks: Left and right hip, left and right ankle<br>Data: Vertical (Y) position in pixels | Median across each one-second interval |
| **Stride Time** | Time between two consecutive heel strikes of the same foot (seconds) | Landmarks: Right and Left Ankle<br>Data Source: Vertical (Y) position in MediaPipe pose units | Median across all strides |
| **Cadence** | Steps per minute | Data Source: Vertical distance between ankles, from stride time calculation | Mean across each walking segment |
| **Stride Width** | Distance between the right and left heel while both are in contact with the ground (cm) | Landmarks: Right and Left Heel<br>Data Source: Vertical (Y) and horizontal (X) positions in meters from MediaPipe world | Median across all strides |

Step and stride parameter definitions follow Huxham et al <sup>25</sup>

### Statistical Analyses

#### Participant Demographics

To describe the study sample, descriptive statistics were used (median (IQR), n (%)).

#### Iterative Phases of Home Video Data Acquisition

To evaluate data acquisition, instructions, adoption, and technical feasibility across Phase I and Phase II, descriptive statistics were used (e.g., percentages of participants who uploaded any videos, usable videos by visual review, and high-quality videos). In visual review after Phase I, usable videos were defined as those with the participant’s full body in frame with no objects occluding the camera view. After video processing, high-quality videos were defined as those with all four gait parameters successfully estimated, indicating sufficient quality for gait analysis.

To evaluate differences in clinical and demographic characteristics among participants who 1) consented to the home video sub-study vs. those who declined or were not approached, and 2) those who uploaded vs. did not upload home videos, Fisher’s exact test and Wilcoxon rank sum tests were used.

#### In-Clinic and Home Video Quality

To further evaluate the feasibility of home video data collection, the quality of videos collected at home by PwMS was compared to those collected in the clinic by study staff (e.g., median (IQR) video and walking segment duration, percentage of high-quality videos with all parameters estimated).

All video analysis steps were conducted in Python 3.10. Statistical analysis was performed using Python 3.10 and R 4.5.2.

## RESULTS

### Participant Demographics and Clinical Characteristics

In-clinic videos were available for 132 PwMS, collected between September 2022 and September 2025; 77 participants had a second consecutive time point. The cohort had low-moderate disability and was representative of individuals living with MS: mean age was 49 years (IQR: 39-60); 70% were female. Median disease duration was 8 years (IQR: 4-15), median EDSS was 2.5 (IQR: 1.4-4.0), and median T25FW was 4.70 seconds (IQR: 3.98-6.10); 77% had relapsing-onset MS (**Table 2**).

**Table 2.** Participant demographic and clinical characteristics.

| Characteristic | In-Clinic Videos<br>N = 132<sup>1</sup> | Home Videos<br>N = 55<sup>1</sup> |
| --- | --- | --- |
| Age (Years) | 49 (39, 60) | 56 (42, 66) |
| Sex |  |  |
| Female | 93 (70%) | 42 (76%) |
| Male | 38 (29%) | 12 (22%) |
| Non-Binary | 1 (0.8%) | 1 (1.8%) |
| Race/Ethnicity |  |  |
| White Non Hispanic | 92 (70%) | 41 (75%) |
| Hispanic or Latino | 12 (9.1%) | 3 (5.5%) |
| Asian | 12 (9.1%) | 4 (7.3%) |
| Black Or African American | 6 (4.5%) | 1 (1.8%) |
| Other/Unknown/Declined | 10 (7.6%) | 6 (11%) |
| Disease Duration (Years) | 8 (4, 15) | 13 (3, 22) |
| MS Subtype |  |  |
| Relapsing-onset | 101 (77%) | 43 (78%) |
| Progressive | 27 (20%) | 11 (20%) |
| CIS (Clinically Isolated Syndrome) | 0 (0%) | 0 (0%) |
| Subtype not specified | 4 (3.0%) | 1 (1.8%) |
| EDSS | 2.5 (1.5, 4.0) | 3.0 (2.0, 4.5) |
| T25FW (Seconds) | 4.70 (3.98, 6.10) | 4.85 (4.15, 6.10) |

<sup>1</sup>n (%); Median (Q1, Q3)

Home videos were available for 55 PwMS at a single timepoint, collected between May 2023 and March 2026. The cohort also had low-moderate disability: mean age was 56 years (IQR: 42-66); 76% were female. Median disease duration was 13 years (IQR: 3-22), median EDSS was 3.0 (IQR: 2.0-4.5), and median T25FW was 4.85 seconds (IQR: 4.15-6.10); 78% had relapsing-onset MS (**Table 2**).

### Iterative Phases of Home Video Data Acquisition

#### Phase I

In the initial phase, the goal was to evaluate an “optimal” video acquisition protocol. Here, participants were asked to record four videos, two in the sagittal plane and two in the frontal plane (**Figure 1B**). While sagittal-plane videos are less user-friendly and require more space, these were considered “optimal” because they facilitate easier estimation of gait parameters related to distance traveled (i.e., step length, velocity). In contrast, frontal-plane videos can be recorded in smaller spaces, allowing data collection in clinical hallways, small apartments, and other more naturalistic settings.

Following visual inspection of home videos from 30 Phase I participants, the Phase II protocol was optimized to address Phase I acquisition pitfalls.

#### Phase II Refinements

##### Acquisition

A visual review of all Phase I videos found that the sagittal videos were of lower quality than the frontal-plane videos. Only 50% of participants uploaded usable sagittal-plane videos upon visual review, with the remaining of poor quality due to difficulty interpreting task instructions or occlusion of key body landmarks from view. In contrast, 87% of participants uploaded usable frontal plane videos (**Figure 4**). Therefore, the protocol was refined to include only frontal plane videos, prioritizing the long-term goal of a user-friendly tool for independent use by PwMS.

**Figure 4.**
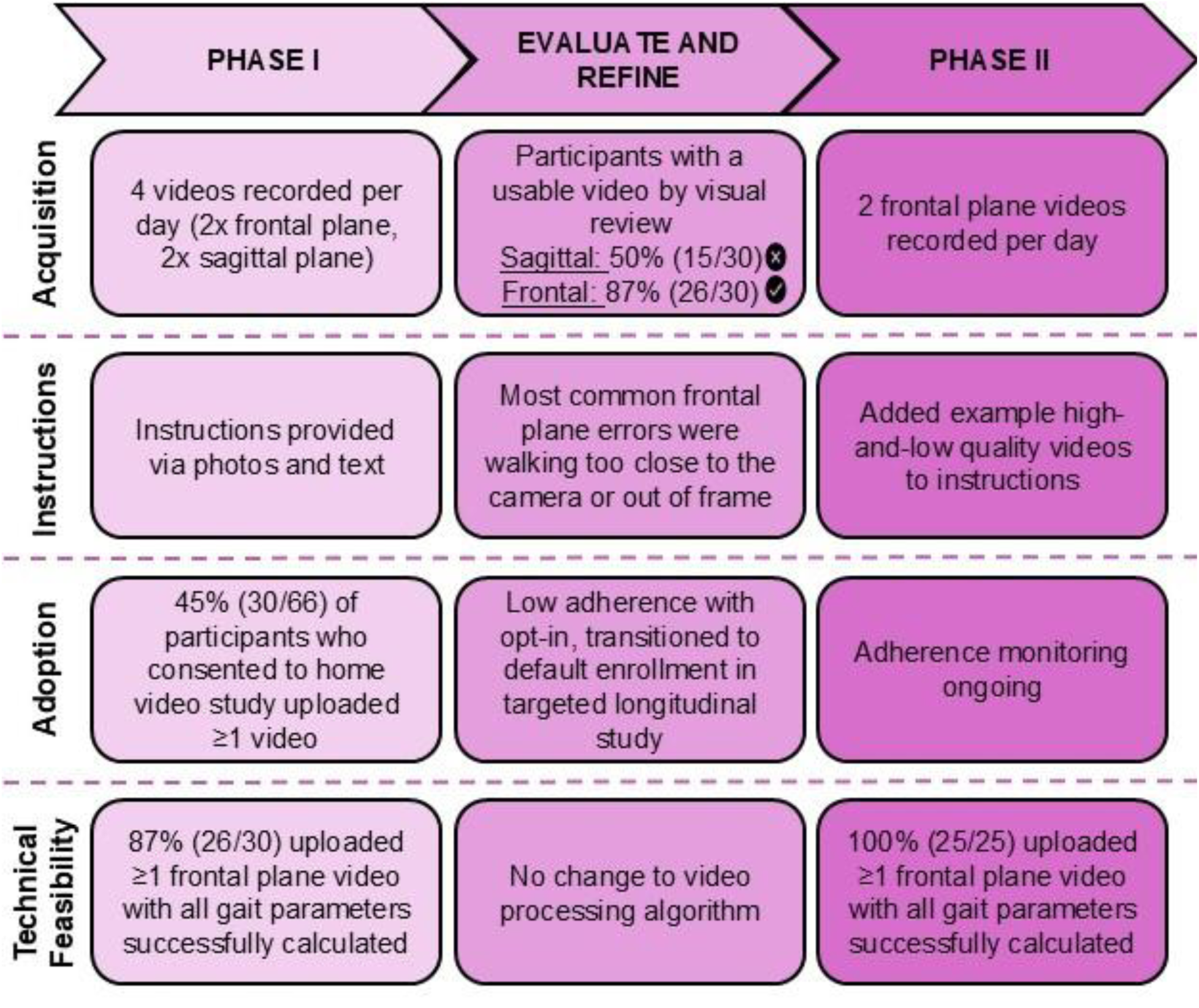
Summary of changes in data acquisition, instructions, adoption, and technical feasibility of home video data collection from Phase I to Phase II.

##### Instructions

In Phase I, the most common limitation in lower-quality frontal-plane videos was participants walking too close to the camera, obscuring lower-body landmarks required to estimate gait parameters. In Phase II, the instructions were updated with examples of low- and high-quality videos. Participants were asked to compare their videos to the examples and re-record if needed.

##### Adoption

In Phase I, all BrainWalk study participants were offered the option to “opt in” to the home videos sub-study in addition to the in-clinic study. This group was overall representative of the PwMS in the larger BrainWalk digital phenotyping study, with no significant differences in demographics or clinical characteristics found between those who consented to home videos (n = 66) and those who either declined or were not approached (n = 112, **Table S1**). Among the 66 who consented to participate, adoption was low, with only 45% (30/66) uploading ≥1 video (**Figure 4**). Those who uploaded home videos had a trend toward longer median disease duration than those who did not (p = 0.049); no other significant differences in demographic or clinical characteristics were found (**Table S2**).

To improve adoption in Phase II, instructions were revised, and video acquisition and upload protocols were simplified. Video collection then became a default component of the BrainWalk study, with ongoing adoption and engagement monitoring.

##### Technical Feasibility

Across both Phase I and II, the majority of participants uploaded high-quality frontal-plane videos sufficient for estimating all gait parameters. The percentage of participants who uploaded a high-quality frontal-plane video increased from Phase I to II. In Phase I, 87% of participants (26/30) uploaded ≥1 high-quality frontal-plane video. This increased to 100% (25/25) in Phase II. All home videos recorded in the frontal plane from both Phase I and II were included in the subsequent analysis.

### In-Clinic and Home Video Quality

A total of 572 frontal plane walking videos were processed. Study team members recorded 467 videos of PwMS at UCSF, and PwMS self-recorded 105 videos at home. In in-clinic videos, participants first completed the PWS task (230 videos), followed by FW (237 videos). All home walking videos were at PWS, with right turns in the first (53 videos) and left turns in the second (52 videos).

Compared with the in-clinic videos, home videos recorded by PwMS were of similar duration and had a similar percentage of high-quality videos.

#### Video and Walking Segment Duration

Of the in-clinic videos, median FW video duration was 22.6 seconds (IQR 9.1), and median PWS video duration was 29.0 seconds (IQR 8.4). Across all frontal-plane home videos, the median video duration was 30.4 seconds (IQR 12.5). Both FW and PWS in-clinic videos had a median of 3 walking segments per video, with FW median walking segment duration of 3.6 seconds (IQR 1.1) and PWS median walking segment duration of 4.4 seconds (IQR 1.5). Similarly, home videos had a median of 4 walking segments, with a median walking segment duration of 3.8 seconds (IQR 1.5).

#### Percentage of High-Quality Videos

Of the in-clinic videos, all four video-derived parameters were estimated from 92% FW (217/237) and 90% PWS videos (206/227) (**Figure S5**). The percentage of home videos of sufficient quality for gait analysis was similar to that of in-clinic videos. All four video-derived parameters could be estimated from 92% of home videos (97/105 overall; 50/53 right turns, 47/52 left turns) (**Table 3, Figure S6**).

**Table 3.**
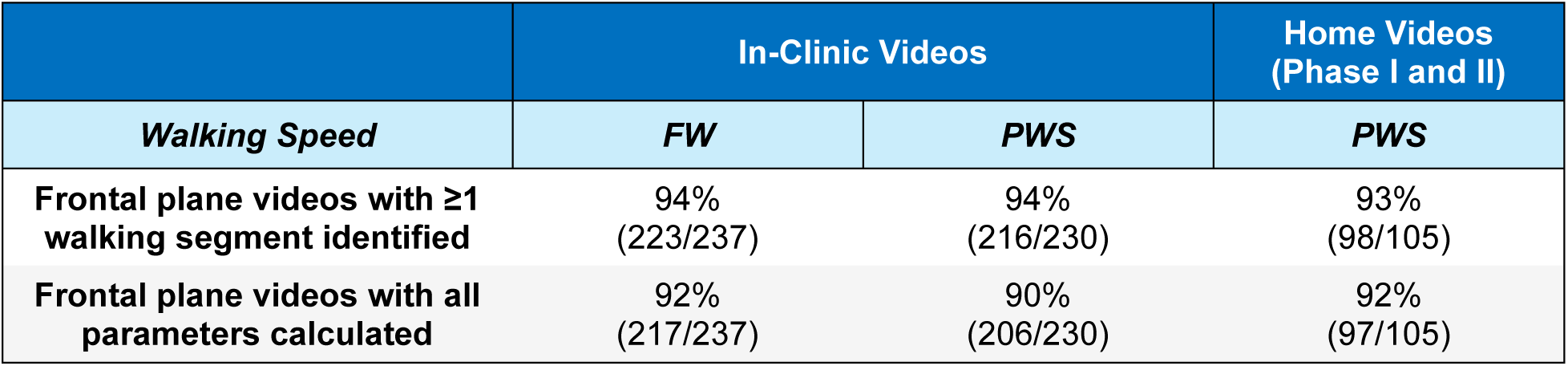
The percentage of high-quality videos was similar between those recorded in-clinic and those recorded by PwMS at home.

|  | In-Clinic Videos |  | Home Videos<br>(Phase I and II) |
| --- | --- | --- | --- |
| *Walking Speed* | *FW* | *PWS* | *PWS* |
| **Frontal plane videos with ≥1 walking segment identified** | 94%<br>(223/237) | 94%<br>(216/230) | 93%<br>(98/105) |
| **Frontal plane videos with all parameters calculated** | 92%<br>(217/237) | 90%<br>(206/230) | 92%<br>(97/105) |

## DISCUSSION

This study aimed to develop a low-burden, accessible method for collecting gait information from PwMS and to evaluate its feasibility by comparing the quality of home-recorded videos with that of in-clinic videos. The pose-estimation-based method described herein estimates spatiotemporal gait parameters from videos recorded with a single frontal-plane camera, expanding the range of possible data-collection settings to include small apartments and hallways. The use of a consumer-grade camera further improves scalability, as PwMS can record these videos using a device they already own (e.g., a smartphone, tablet, or laptop). With appropriate validation, gait parameters such as slower gait speed, decreased cadence, and longer stride times^18^ from consumer-grade devices could provide a low-cost method for monitoring gait-related disability progression over time (**Figure S7**).

This study identified several modifications that improved the quality of patient-uploaded videos. Primarily, to prioritize ease of recording, we removed the requirement to record sagittal-plane videos and focused only on user-friendly frontal-plane videos. The instructions to participants were also updated to include example high- and low-quality videos. Using these updated instructions, 100% of participants in Phase II uploaded at least one video that was of high enough quality (e.g., full body in frame, sufficient walking duration) for gait analysis. When evaluating all frontal-plane videos, we found that the percentage of high-quality videos was similar between those recorded in-clinic by the study team and those recorded at home by PwMS (>90%), highlighting the feasibility of the home video data collection approach.

Participants’ adoption of the walking video protocol differed from our prior work on hand function, in which PwMS self-recorded themselves seated while performing tasks such as buttoning a shirt.^12^ In that study, of 50 consented, 44 (88%) uploaded baseline videos. This difference in adoption could be due to the relative ease and familiarity of seated dexterity tasks (less camera positioning) compared with the greater learning for gait tasks. Technically, the gait videos required technological modifications to the electronic video upload solution to accommodate larger file sizes. Ongoing monitoring of adoption and engagement will occur during the longitudinal home video collection protocol to ensure improvements in response to such changes.

Study limitations include the low-to-moderate disability cohort enrolled. Future work will evaluate the feasibility among participants with greater disability due to MS, and stakeholder feedback interviews will be utilized to identify barriers to use across a wider range of MS impairment. Additionally, all home videos were recorded at the participant’s self-selected PWS to prioritize safety, precluding direct comparison with the gold-standard T25FW.^20^ Screening for fall risk via a physical therapy screen, or if unavailable; asking about prior fall history^21^ or subjective questionnaire thresholds (i.e.,12-item MS Walking Scale [MSWS-12] ≥ 46%)^22–24^ will help determine which patients can safely record fast-walking videos at home going forward, prompting focused feasibility analyses of FW home video collection.

This study demonstrates the initial feasibility of extracting gait data from videos self-recorded by PwMS in their own homes. This novel gait analysis method, using a single consumer-grade video and open-source software, could provide a quantitative snapshot of patient gait at routine intervals, supporting low-burden, objective assessments of changes in response to treatment and rehabilitation interventions. Future validation efforts will explore the cross-sectional and longitudinal validity of video-derived gait parameters relative to standard outcome measures (e.g., EDSS,^14^ T25FW^20^).

## Statements and Declarations

### Data Availability

The data supporting these findings of this study are available upon request from the study author. The data are not publicly available due to privacy or ethical restrictions.

## Author Contributions

MM conceptualized, designed the study, conducted statistical analyses, designed figures, and wrote the first draft. RB provided supervision, conceptualized the study, and acquired funding. AT-E, MJM, VJB, and RB contributed to study design, analysis plan, and data interpretation. MM and YA developed the algorithm to estimate gait parameters. SP curated study data. AC, NS, JW, KH, and JB collected and curated the data. All authors contributed to review and editing.

## Acknowledgements

The authors would like to thank the patients who participated in this study. This study was funded by F. Hoffmann-La Roche Ltd., Basel, Switzerland as part of the Integrative Neuroscience Collaborations Network (Bove, PI).

