## Supplemental Material for "Feasibility of Patient-Uploaded Videos for Gait Assessment in Multiple Sclerosis"

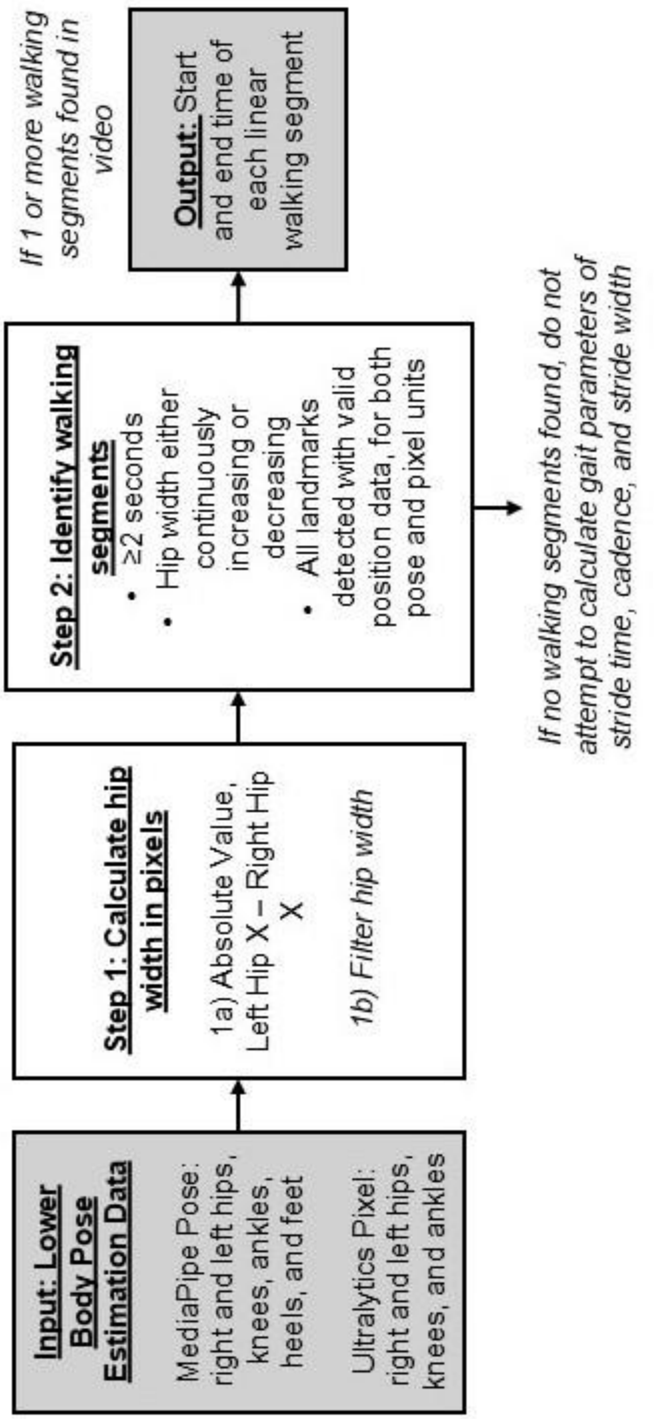


**Supplemental Figure 1. Steps of the algorithm to identify walking segments.**

### Video-Derived Gait Parameters

*Velocity by Pixels*

A measure of velocity was challenging to extract given the video acquisition parameters, namely the recording in the frontal plane and the lack of reference distances. To approximate walking velocity, the change in pixel size of the same object as a person moved toward and away from the camera was calculated. This proxy measure evaluates the vertical distance between two landmarks, in pixels, per second. While it is not a direct substitute for true velocity, a greater change in pixel distance per second was expected for a person walking quickly relative to someone walking slower.

To calculate this parameter, the right and left hip and ankle vertical (Y) position data in pixels were smoothed with the rolling mean of each window of 10 frames. Next, the absolute vertical distance between the right hip and right ankle and the absolute vertical distance between the left hip and left ankle were calculated. The average of the right hip to ankle distance and left hip to ankle distance was calculated at each frame, and the averaged hip-to-ankle distance time series data were then smoothed with a rolling mean over each window of 25 frames.

The averaged hip-to-ankle data were then grouped into one-second intervals. For each interval, the change in hip-to-ankle distance from the last to the first frame of each second was calculated (**Figure S2**). This change was then divided by the hip-to-ankle distance in the first frame of the current one-second increment to account for a larger change in pixel height for the same distance traveled when the person was closer to the camera. One-second intervals that contained either a local minimum, local maximum, or any gaps with missing landmark position data were excluded from the analysis to avoid calculating this parameter over sections of the video with either noisy pose estimation data or when the individual turned. The median change in pixel hip-to-ankle vertical distance over all the included one-second increments was reported as a summary measure for each video.

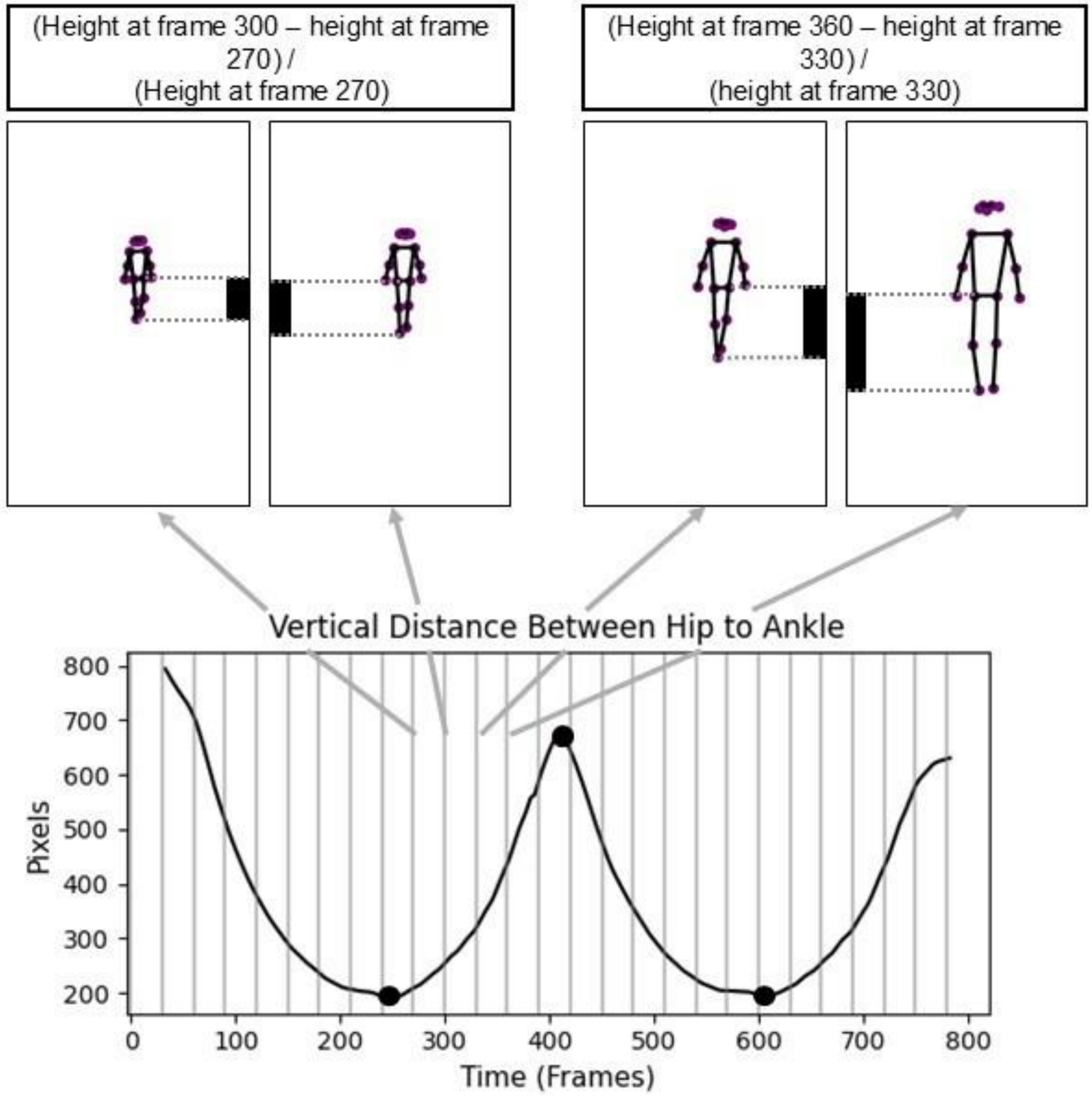


**Supplemental Figure 2. Change in pixel height per second as a measure of velocity.** *Plot of the vertical distance between the hip and ankle in pixels over a subsection of a walking video recorded at 30 frames per second. UltralyticsYOLOv8-pose landmark positions with the calculation for the change in pixel height from the 270$^{th}$ to the 300$^{th}$ frame and the 330$^{th}$ to the 360$^{th}$ frame. The change in pixel height per second was divided by the pixel height at the first frame of the current second to account for the larger changes in pixel height when the individual was closer to the camera, as shown in the difference between the 270$^{th}$ and 300th frames and the 330th and 360th.*

*Stride Time*

To calculate the time between two consecutive heel strikes of the same foot, gaps in the vertical position of the left and right ankle MediaPipe pose landmarks up to 1.2 seconds were filled using linear interpolation. The vertical distance between the right and left ankle was calculated and smoothed with a rolling mean over every 15 frames. The time between each ankle vertical distance local maxima were the stride times of one foot. The time between each ankle vertical distance local minima were the stride times of the other foot (**Figure S3)**, with modified calculations from Stenum et al[1]). If both the right and left foot had one or fewer strides identified, stride time was not calculated from that walking segment.

The stride time for every stride in each linear walking segment was calculated. Summary measures, including mean, median, and standard deviation, of these strides were reported for each video.

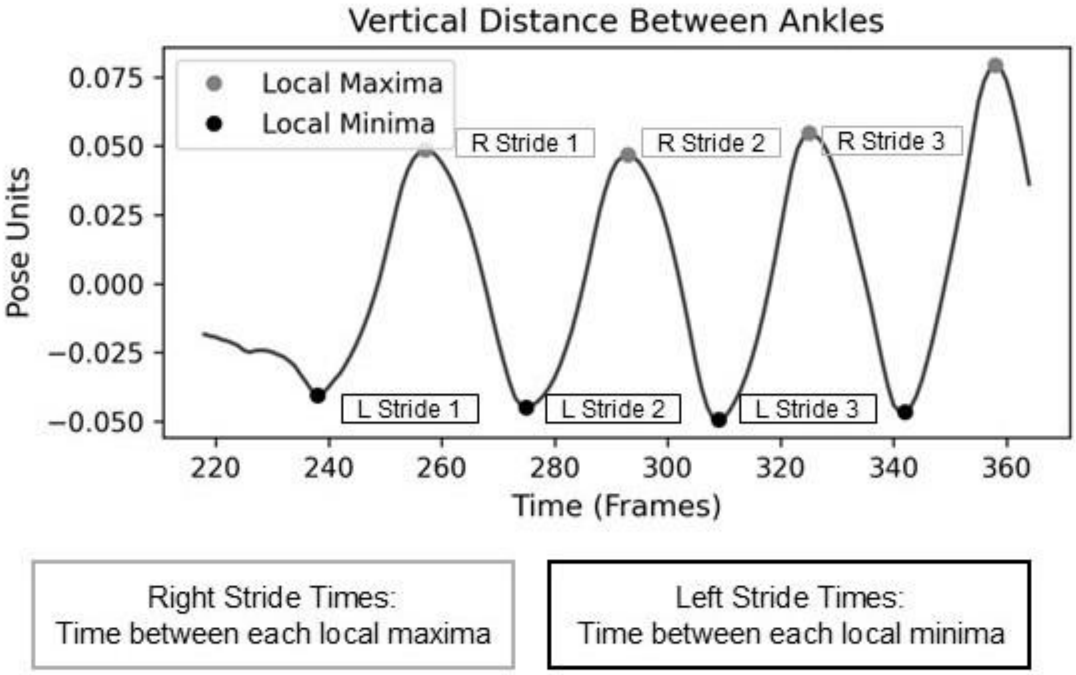


**Supplemental Figure 3. Estimation of stride time.**
*Plot of the vertical distance between the right and left ankle for a single walking segment of a video recorded at 30 frames per second. Local maxima were used to calculate stride time for the right foot and local minima were used to calculate stride time for the left foot.*

*Cadence*

The total number of local minima and maxima of the vertical distance between the right and left ankle from the stride length calculation were added together to estimate the total number of steps per walking segment. Cadence for each walking segment was calculated as the total number of steps plus one divided by the walking segment time in seconds (steps/second). This value was then multiplied by 60 to convert to steps/minute. The mean cadence across all walking segments was reported for each video.

Similar to stride time, if both the right and left foot only had one or zero strides identified, cadence was not calculated for that walking segment.

*Stride Width*

To calculate the distance between the right and left heel while both are in contact with the ground, gaps in the MediaPipe world heel data up to 1.2 seconds were filled using linear interpolation (both ankles, X and Y axis). The vertical distance between the heels was calculated and smoothed with a rolling mean over every 15 frames. The frames where the vertical distance between heels crosses zero were identified to estimate when both heels are on the ground (**Figure S4)**. The width of each stride was calculated as the horizontal (X) distance between the heels at these zero-crossing frames. Lastly, the MediaPipe world output of meters was converted to centimeters.

The stride width for every stride in each linear walking segment was calculated. Summary measures, including mean, median, and standard deviation, of these strides were reported for each video.

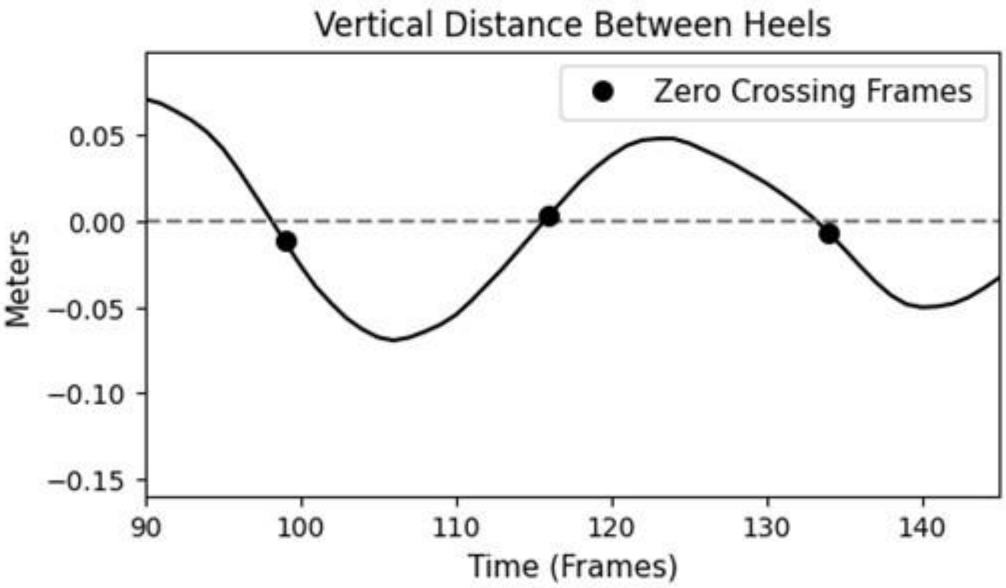


**Supplemental Figure 4. Estimation of stride width.**
*Plot of the vertical distance between heels in meters from a subsection of a walking video recorded at 30 frames per second. The frames in which the vertical distance between the heels crossed zero indicated that both heels were on or near the ground, as shown by black dots. Stride width was estimated by calculating the horizontal (X) distance between the heels at each of these frames.*

**Supplemental Table 1. Demographics and clinical characteristics of participants who consented to home videos versus those that did not in Phase I.**

| Characteristics | Declined/Not Approached: BW Only N = 112[1] | Consented N = 66[1] | p-value[2] |
|---|---|---|---|
| Age (Years) | 48 (38, 61) | 48 (37, 59) | 0.7 |
| Sex | | | 0.5 |
| Female | 79 (71%) | 48 (73%) | |
| Male | 33 (29%) | 17 (26%) | |
| Non-Binary | 0 (0%) | 1 (1.5%) | |
| Race/Ethnicity | | | 0.6 |
| White Non Hispanic | 77 (69%) | 42 (64%) | |
| Hispanic or Latino | 13 (12%) | 7 (11%) | |
| Asian | 6 (5.4%) | 7 (11%) | |
| Black Or African American | 8 (7.1%) | 3 (4.5%) | |
| Other/Unknown/Declined | 8 (7.1%) | 7 (11%) | |
| Disease Duration (Years) | 8 (4, 15) | 7 (3, 13) | 0.2 |
| MS Subtype | | | 0.7 |
| Relapsing-onset | 81 (72%) | 52 (79%) | |
| Progressive | 28 (25%) | 13 (20%) | |
| CIS (Clinically Isolated Syndrome) | 0 (0%) | 0 (0%) | |
| Subtype not specified | 3 (2.7%) | 1 (1.5%) | |
| EDSS | 2.5 (1.5, 4.0) | 3.0 (2.0, 4.5) | 0.064 |
| T25FW (Seconds) | 4.63 (3.98, 6.10) | 5.00 (4.15, 6.10) | 0.3 |

[1]n (%); Median (Q1, Q3)

[2]Fisher's exact test; Wilcoxon rank sum test

**Supplemental Table 2. Demographics and clinical characteristics of participants who consented and uploaded videos in Phase I vs. those who did not upload any videos.**

| Characteristics | Did Not Upload Videos N = 36[1] | Uploaded Videos N = 30[1] | p-value[2] |
|---|---|---|---|
| Age (Years) | 46 (36, 57) | 51 (38, 63) | 0.5 |
| Sex | | | 0.4 |
| Female | 24 (67%) | 24 (80%) | |
| Male | 11 (31%) | 6 (20%) | |
| Non-Binary | 1 (2.8%) | 0 (0%) | |
| Race/Ethnicity | | | 0.3 |
| White Non Hispanic | 19 (53%) | 23 (77%) | |
| Hispanic or Latino | 6 (17%) | 1 (3.3%) | |
| Asian | 5 (14%) | 2 (6.7%) | |
| Black Or African American | 2 (5.6%) | 1 (3.3%) | |
| Other/Unknown/Declined | 4 (11%) | 3 (10%) | |
| Disease Duration (Years) | 6 (1, 10) | 9 (4, 16) | 0.049 |
| MS Subtype | | | 0.5 |
| Relapsing-onset | 28 (78%) | 24 (80%) | |
| Progressive | 8 (22%) | 5 (17%) | |
| CIS (Clinically Isolated Syndrome) | 0 (0%) | 0 (0%) | |
| Subtype not specified | 0 (0%) | 1 (3.3%) | |
| EDSS | 2.5 (2.0, 4.0) | 3.0 (2.0, 6.0) | 0.4 |
| T25FW (Seconds) | 5.03 (4.28, 6.15) | 4.98 (4.15, 6.10) | 0.8 |

[1]n (%); Median (Q1, Q3)

[2]Fisher's exact test; Wilcoxon rank sum test

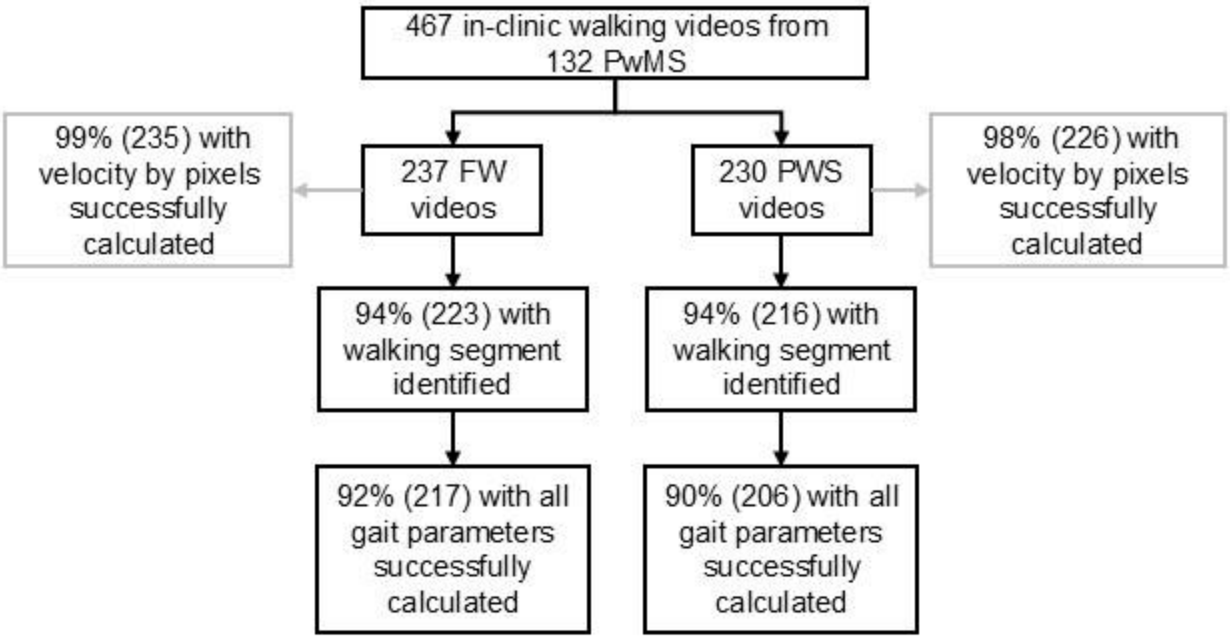


**Supplemental Figure 5. Preferred walking speed (PWS) and fast walking (FW) in-clinic videos with walking segments and all video-derived gait parameters extracted.**

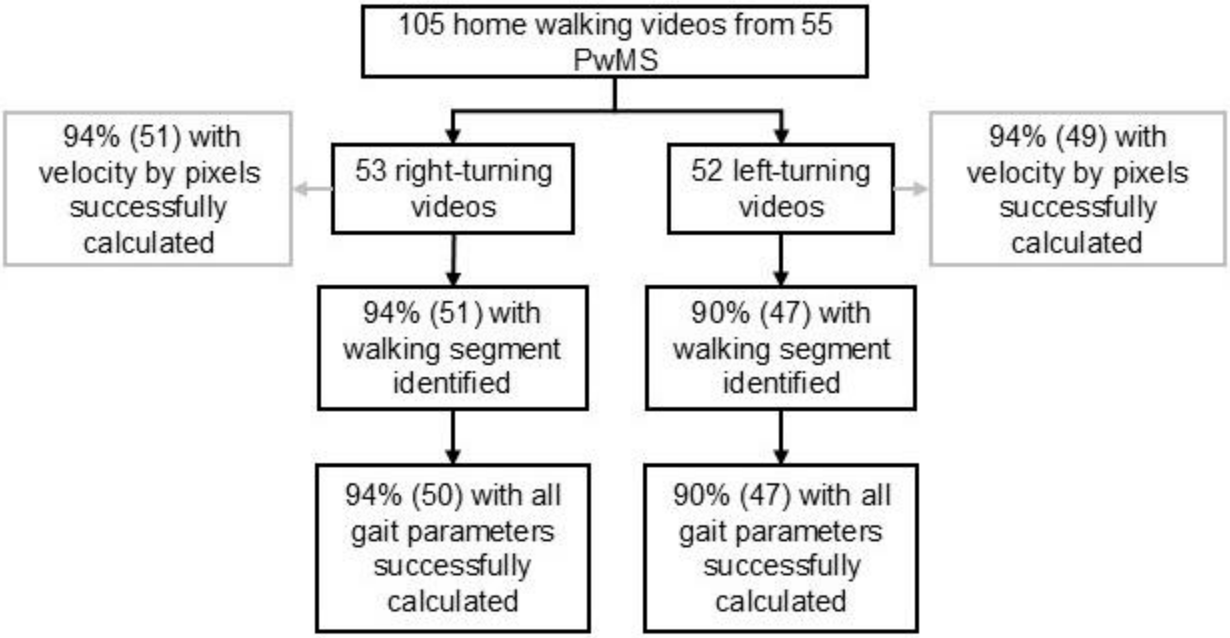


**Supplemental Figure 6. Home videos with walking segments and all video-derived gait parameters extracted.**

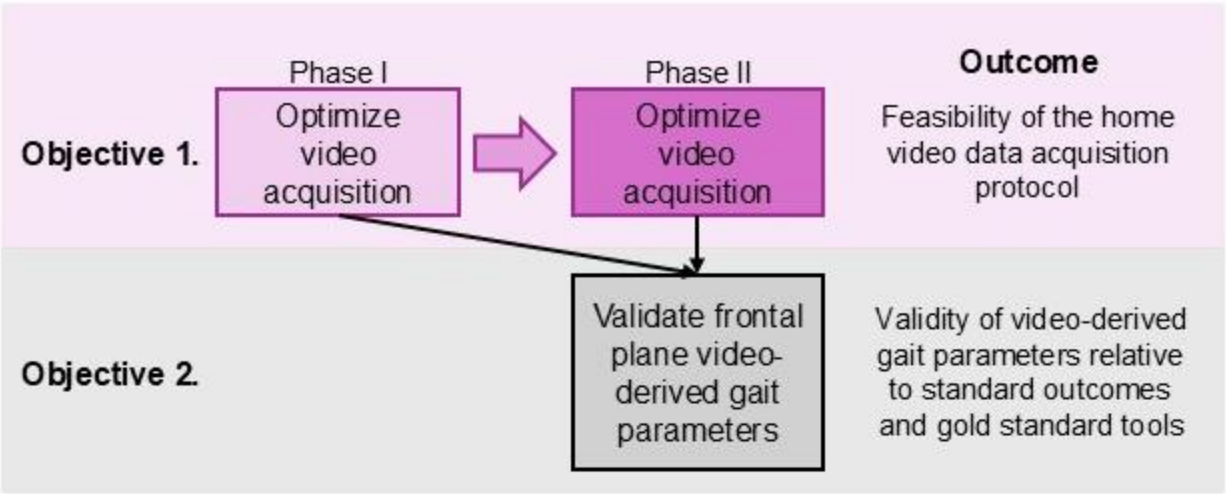


**Supplemental Figure 7. Schema of study objectives.**

*This work focused on objective 1, with the main objective of evaluating the feasibility of home video data collection and optimizing data acquisition methods. Future work will aim to achieve objective 2 and to validate the gait parameters estimated from frontal-plane videos.*
